# Hemoglobin specific volume width promotes the prevalence and poor long-term prognosis of American adult hypertensive patients: the NHANES 1999-2020

**DOI:** 10.1101/2023.08.15.23294146

**Authors:** Lin Zhang, Jun Chang, Kaiyue Wang, Ying Zhou, Liming Wang, Lele Zhang, Xuemei Zhang, Zhifei Fu, Lifeng Han, Xiumei Gao

## Abstract

**Background:** Hypertensive patients are always accompanied by erythrocyte dysfunction. However, current erythrocyte-related indicators can’t explain hypertension’s prevalence and long-term prognosis. Therefore, hemoglobin specific volume width (HSW) was first created to explain this phenomenon.

**Methods:** 59,867 adult participants from National Health and Nutrition Examination Survey (NMAHES) were included. HSW’s quartiles were determined with Q1 [1.88,3.64] cL/g, Q2 (3.64,3.84] cL/g, Q3 (3.84,4.11] cL/g, and Q4 (4.11,11.74] cL/g. 21,006 hypertensive patients had a whole following time 97 (51, 151) months, 15,519 hypertensive patients were alive, and 5,487 were dead. The relationship between HSW and hypertension was analyzed.

**Results:** Among Controls *n*=35,677 and Hypertensive patients *n*=24,190, the percentages of hypertension in quartiles of HSW (Q1, Q2, Q3, and Q4) were 28.59%, 33.35%, 39.37%, and 47.74%. Adjusted odds ratio (OR) in HSW was still significant, 1.23 (95% CI 1.11,1.36). Among dead (*n*=5,487) and alive hypertensive patients (*n*=15,519), the percentages of hypertensive mortality in quartiles of HSW were 17.66%, 20.46%, 20.78%, and 25.02%. The adjusted HSW hazard ratio (HR) was 1.91(95%CI 1.69,2.16). Processing Q1 as reference, the HR for Q4 was 2.35 (95% CI 2.06, 2.69). Males had a higher risk (HR: 1.53 95% CI 1.24,1.89) of poor prognosis than females (HR: 1.48 95% CI 1.17,1.87). Individuals <=60 years old (HR: 2.25 95% CI 1.78,2.85) had a higher risk of poor prognosis than those >60. Hypertensive patients with HSW > 3.89 cL/g had a poor prognosis than HSW <= 3.89 cL/g.

**Conclusions:** HSW is an innovative independent risk factor for hypertensive prevalence and long-term prognosis.

## 1. Introduction

Red blood cell (RBC) damage is a recognized feature of hypertensive patients, including reduced RBC membrane fluidity ^1^ and impaired RBC deformability ^2,3^. The reduced RBC membrane fluidity might participate in the pathogenesis of hypertension^4^. However, few direct evidence verified a causal relationship between RBC-related markers for hypertension’s prevalence or long-term prognosis. For example, hemoglobin concentration was positively associated with blood pressure ^5^ but could not promote the development of hypertension ^6^. Red blood cell distribution width (RDW), an index related to erythrocyte homeostasis ^2,7^, might trigger the evolution of hypertension ^8^. But absent evidence confirmed the predictive value of RDW in hypertensive patients. Although erythrocyte dysfunction is a clinical feature of hypertensive patients, current erythrocyte-related indexes do not explain hypertension’s prevalence and long-term prognosis.

Hemoglobin specific volume width (HSW), first proposed and developed in this work, is a novel RBC-related measure that mirrors the volume fluctuation of RBC corresponding to every unit of hemoglobin. HSW is similar but different to RDW, which is a variation in RBC volume ^9^. And HSW emphasizes the variability volume fluctuation of RBC corresponding to every unit of hemoglobin. However, previous research work had not focused on HSW in any disease. And the calculation strategy of HSW was developed for the first time in this work. We hypothesized that there was a specific association between HSW and hypertensive patients, so relevant experiments were designed to verify it.

Three objectives are in this study: i) analyze the prevalence between HSW in Control and hypertensive patients; ii) explore the optimal value of HSW to define the prognosis of hypertensive patients; iii) analyze the prognosis of hypertensive patients at various levels of HSW.

## 2. Methods

### 2.1 Calculation of HSW

As the inverse of density, the specific volume describes the volume per unit mass. It is widely applied in muscle testing ^10^ and food ^11–13^ but never in hemoglobin. For example, the muscle specific volume illustrates the skeletal muscle volume in different individuals ^10^. In the food field, the bread specific volume width indicated the fluctuation of the expanded bread volume with the same mass ^14,15^ at various conditions.

Similarly, the HSW can reflect fluctuation in the volume of RBC with the same hemoglobin mass. Previous research had not focused on the HSW in any disease, and we first proposed it. So how to measure or calculate the HSW?

As the name implies, the mean corpuscular haemoglobin concentration (MCHC) ^16^ is the mean value of hemoglobin density in red blood cells rather than whole blood. Because specific volume is the inverse of density, the hemoglobin specific volume can be calculated by MCHC. However, hemoglobin specific volume and HSW are different in that the former emphasizes the specific volume, while the latter focuses more on the fluctuation of the specific volume. So the RBC volumetric fluctuation parameter needs to be taken into account. Interestingly, the RDW ^17^ was used to describe the variability or fluctuation of red blood cell volume. That is, the HSW can be calculated with RDW and HSV. In summary, the rude calculation strategy of HSW is finally formulated, and HSW is equal to RDW divided by MCHC. While the calculation strategy of HSW was developed, more details are needed to justify it.

So, the calculated detail of HSW is shown in **Fig. 1**. First, RDW equals erythrocyte volume’s standard deviation (EV-SD) by the mean corpuscular volume (MCV) ^18^. Second, the MCHC ^19^ equals mean cell hemoglobin (MCH) divided by the mean corpuscular volume (MCV). And HSW equals RDW divided by MCHC. With the process of **Fig. 1**, the HSW is finally equal to EV-SD divided by MCH. From the formula, HSW can characterize fluctuations in red blood cell volume per gram of hemoglobin.

**Fig. 1.**
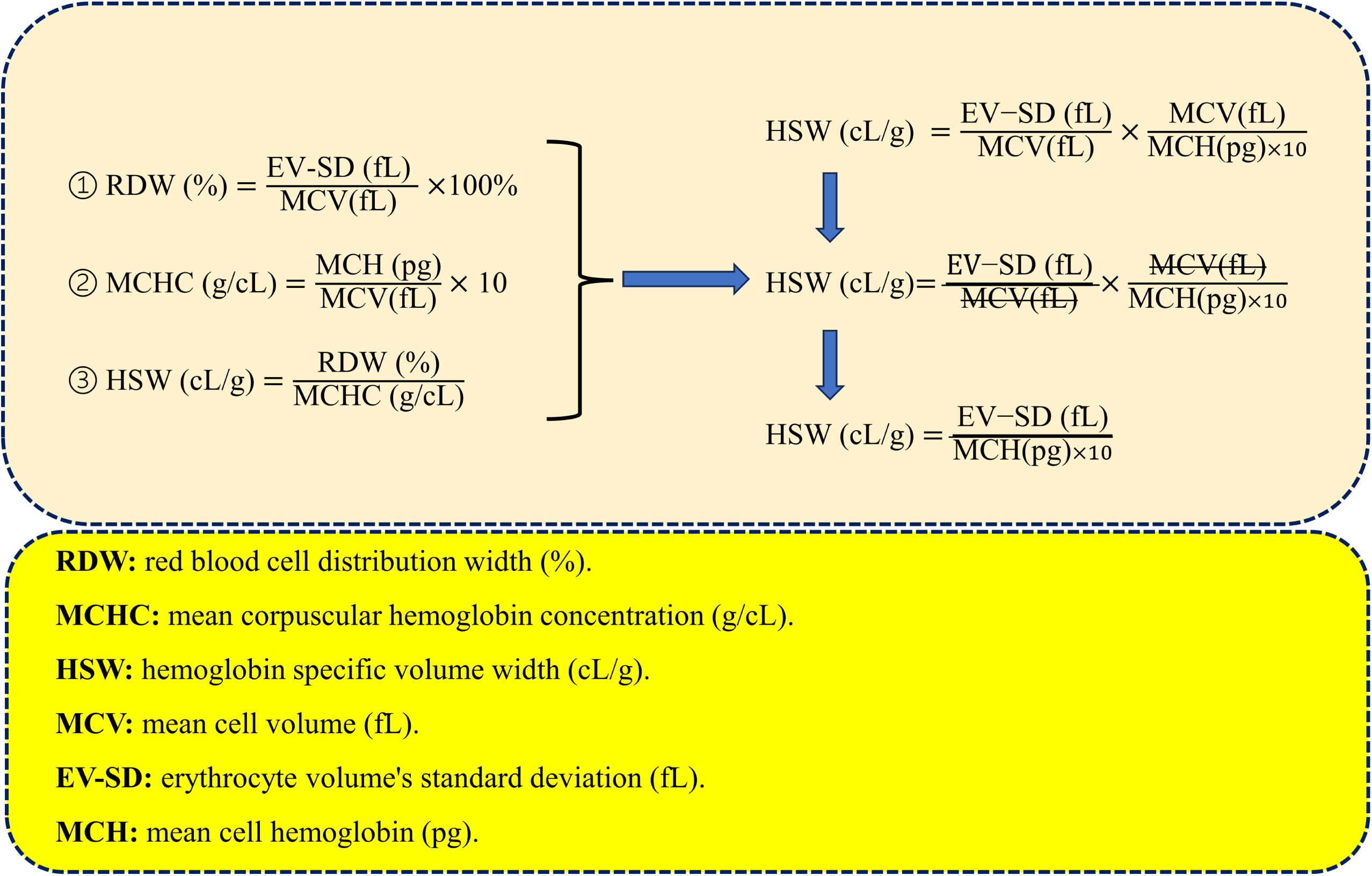
The calculated detail of HSW.

### 2.2 Participants

National Health and Nutrition Examination Survey (NHANES) 1999–2020 offer the primary data. NHANES, as a periodic health-related survey in America, currently contained 116,876 patients and was open to the public free of charge (www.cdc.gov/nchs/nhanes/irba98.htm.). All the research participants provided written informed consent.

In NHANES, the inclusion criteria included age >18 years, HSW presence, blood pressure measurements, and completing lab tests. The following were excluded: absence of the RDW or MCHC (*n*=21,794), missing the hypertensive questionnaire or blood pressure records (*n*=15,001), and age < =18 years (*n*=20,214). Therefore, a total of 59,867 participants were included in the following process. Of the 59,867 participants, 24,190 hypertension patients were specifically filtered for prognosis analysis. And the excluded 3,184 hypertension patients without follow-up time. Finally, 59,867 participants were included for prevalence analysis, and 21,006 hypertension patients were included in the prognosis process (**Fig. 2**).

**Fig. 2.**
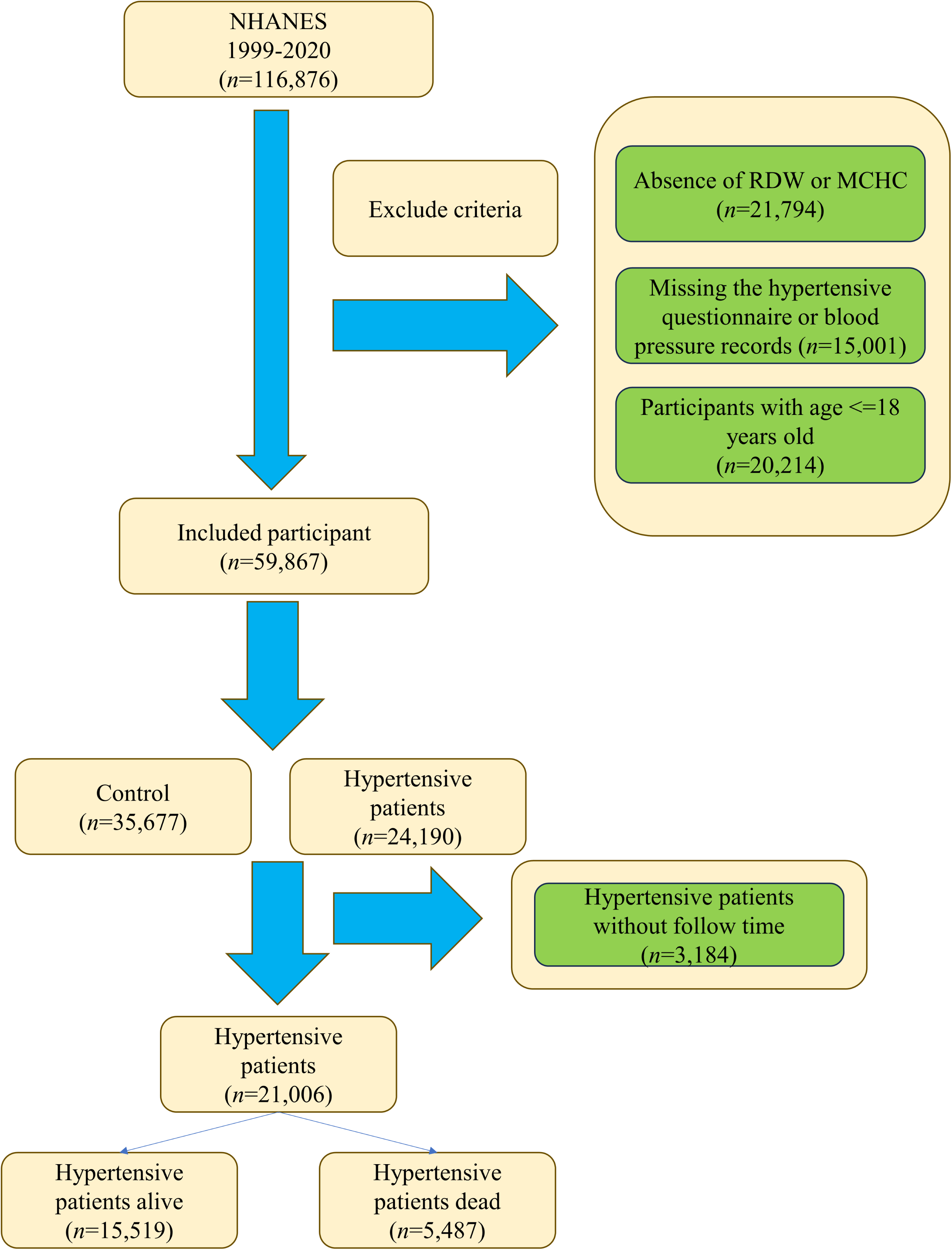
The flow chart selection from the NHANES 1999-2020.

### 2.3 The definition of hypertension

The medical conditions questionnaire (MCQ) mainly obtained the relevant diagnoses (https://wwwn.cdc.gov/nchs/nhanes/default.aspx). The definition of hypertension in this study included three criteria, i) participants answer the MCQ code of bpq020, bpq030, and mcq100; ii) blood pressure measure with 3 times and average systolic blood pressure (SBP) > 140 mmHg or average diastolic blood pressure (DBP) > 90mmHg; iii) related drugs (e.g., nifedipine, valsartan, etc).

### 2.4 Covariates

Several covariates were filtered based on the following criteria, i) demographic variables; ii) characteristics affecting hypertension reported in previous studies; and iii) other variables derived from the clinical experience.

Previous research emphasized several hypertensive diseases, including chronic obstructive pulmonary disease (COPD) ^20,21^, diabetes mellitus (DM) ^22^, and anaemia^23^. COPD was diagnosed with four criteria, FEV1/FVC < 0.7 in post-bronchodilator, the MCQ coded of mcq160g and mcq160p that ever told you had emphysema, age above 40 with a smoke history or chronic respiratory diseottom, and the history of COPD drugs (*e,g.* selective phosphodiesterase-4 inhibitors, mast cell stabilizers, and leukotriene modifiers). The diagnosis of DM includes five criteria, including glycohemoglobin (%) >= 6.5, fasting glucose (mmol/L) >= 7.0, random blood glucose (mmol/L) >= 11.1, two-hour oral glucose tolerance test blood glucose (mmol/L) >= 11.1, and history of drug (*e,g.* diabetes medication and insulin). Impaired fasting glycaemia and impaired glucose tolerance were divided into the preDM group. The Anemia ^24^of adult was defined in adult with four levels, non-Anaemia (female with haemoglobin >=120 g/L or male >=130 g/L), Mild (female with haemoglobin between 110-119 g/L or male 110-129 g/L), Moderate (haemoglobin between 80-109 g/L), and Severe (haemoglobin < 80 g/L).

In addition to the above covariates, other lab tests were tallied, including white blood cell (WBC, 1000 cells/μL), lymphocyte percentage (LymP, %), monocyte percentage (MonP, %), segmented neutrophils percentage (SegneP, %), eosinophils percentage (EoP, %), basophils percentage (BaP, %), segmented neutrophils number percentage (SeneP, %), eosinophils number count (Eo, 1000 cells/μL), red blood cell count (RBC, million cells/μL), hemoglobin (Hg, g/dl), hematocrit (Hem, %), mean cell volume (MCV, fL), mean cell hemoglobin (MCH, pg), mean cell hemoglobin concentration (MCHC, g/cL), red cell distribution width (RDW, %), platelet count (Plt, 1000 cells/μL), mean platelet volume (MPV, fL), alanine transaminase (ALT, U/L), aspartate transaminase (AST, U/L), total calcium (Ca, mg/dL), total cholesterol (TC, mg/dL), bicarbonate (HCO3, mmol/L), gammaglutamyl transferase (GGT, U/L), total protein (TP, g/dL), triglycerides (TG, mg/dL), uric acid (UA, mg/dL), sodium (Na, mmol/L), and lymphocyte count (Lym, 1000 cells/μL),.

### 2.5 Hypertensive mortality

For NHANES, National Death Index (https://www.cdc.gov/nchs/ndi/index.htm) was linked with NHANES and directly provided the mortality-related data. As previously reported ^25^, hypertensive all-cause mortality information was derived and matched with unique individuals’ IDs. Followup time was defined as the period of blood draws time to death or December 31, 2019.

### 2.6 Statistical analyses

All analysis processes were finished with R software (version 4.2.2). In cleaning clinical data, the R packages *nhanesR* (version 0.9.4.1) were served for NHANES.

In the analysis of variance, if the continuous variables were distributed with Gaussian, the Student’s t-test process, and else Mann–Whitney U. A chi-square test was performed to factor data (e.g., Sex and ethnicity). The continuous variables were presented as means ± SDs and proportions for factor variables.

Logistic regression was developed to explore the odds ratio (OR) in the Control and Hypertension groups. Cox regression was applied to the HR and 95% confidence intervals (CIs) of HSW for hypertensive all-cause mortality. Time-to-death among groups is presented with Kaplan–Meier (KM) curves. Importantly, to optimize the cut-off for HSW in hypertensive patient mortality, restricted cubic spline (RCS) ^26^ was applied in this work. The RCS model (with 3-8 knots) examined the optimal HSW value association with mortality. Only a *P*-value <0.05 was considered statistically significant.

## 3. Result

### 3.1 General characteristics

Among 59,867 participants were included, 31,010 (51.80%) females and 28,857 (48.20%) males, with an average age of 49, and 18,690 (31.22%) were over 60 years old. All the participants were classified into Control (*n*=35,677) or Hypertension (*n*=24,190) individuals, which could represent 213,532,107 American adults (**Table 1**) according to the calculated weights. The HSW level in the Hypertension groups (3.94±0.01) is higher than the Control group (3.83±0.01). Among the 35 characteristics, only two had no significance, including Lym and Na.

**Table 1.**
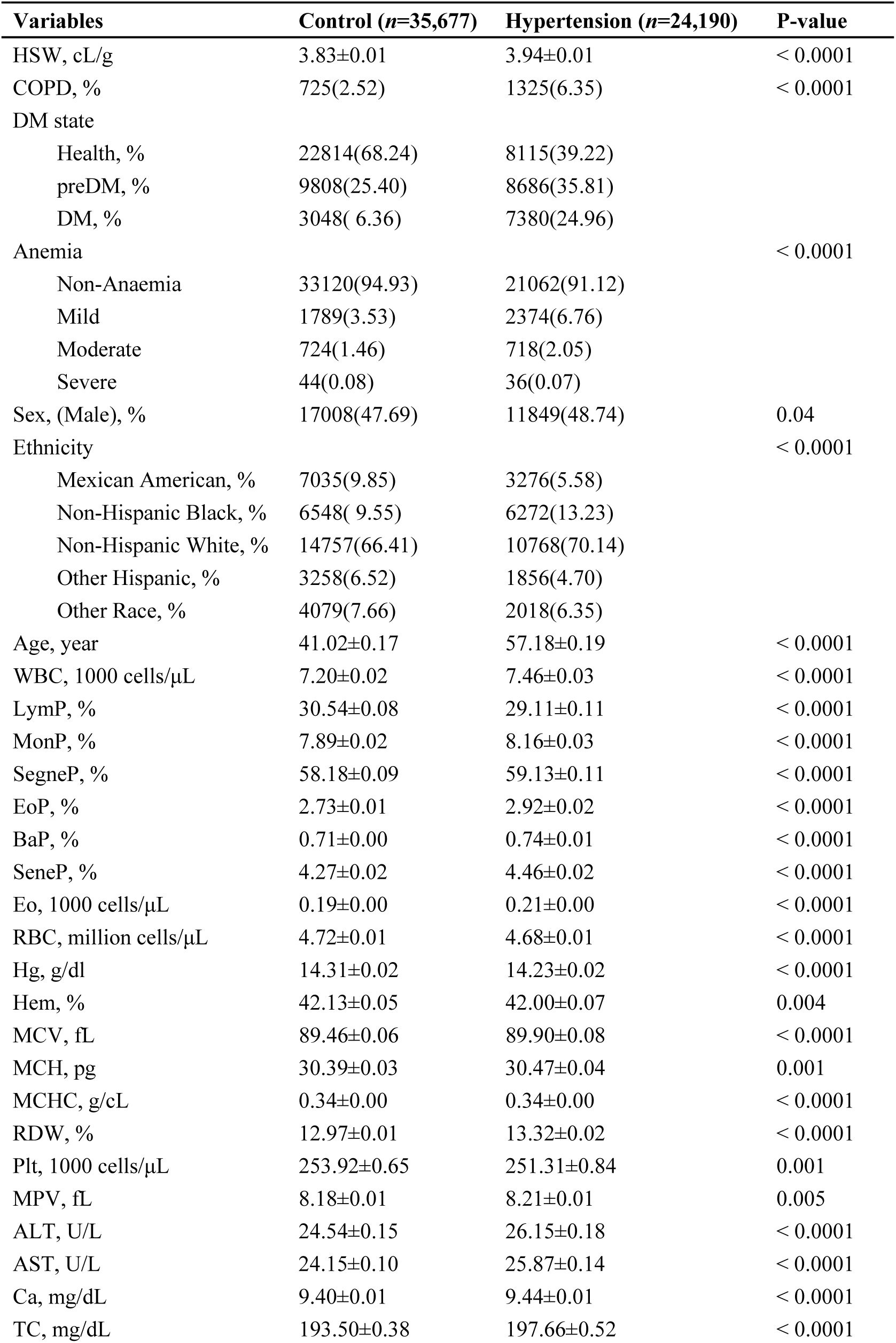
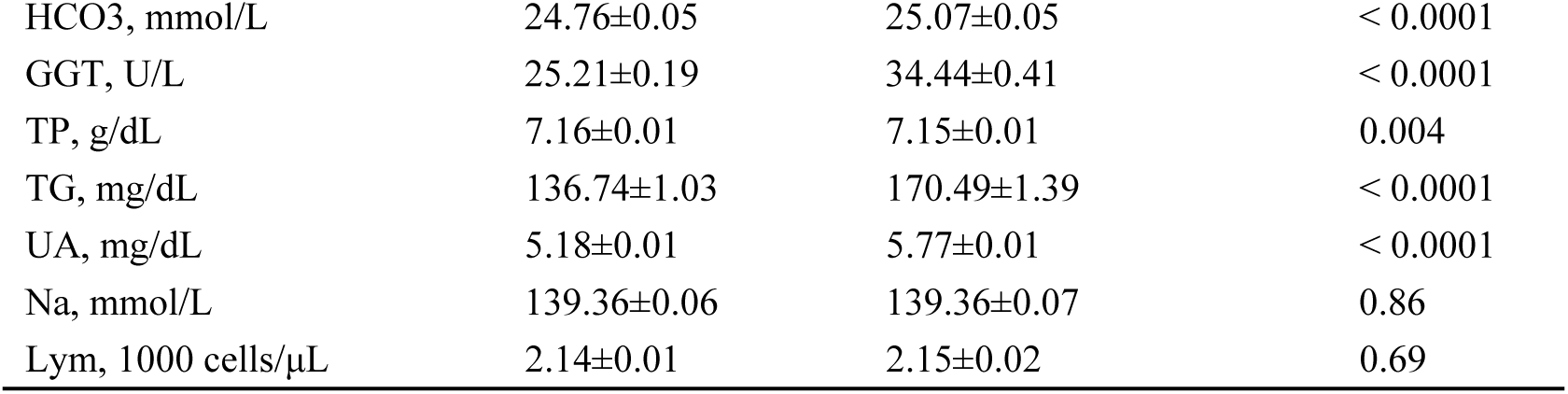
General characteristics for Control and Hypertension patients in the NHANES.

| Variables | Control (n=35,677) | Hypertension (n=24,190) | P-value |
| --- | --- | --- | --- |
| HSW, cL/g | 3.83±0.01 | 3.94±0.01 | < 0.0001 |
| COPD, % | 725(2.52) | 1325(6.35) | < 0.0001 |
| DM state |  |  |  |
| Health, % | 22814(68.24) | 8115(39.22) |  |
| preDM, % | 9808(25.40) | 8686(35.81) |  |
| DM, % | 3048( 6.36) | 7380(24.96) |  |
| Anemia |  |  | < 0.0001 |
| Non-Anaemia | 33120(94.93) | 21062(91.12) |  |
| Mild | 1789(3.53) | 2374(6.76) |  |
| Moderate | 724(1.46) | 718(2.05) |  |
| Severe | 44(0.08) | 36(0.07) |  |
| Sex, (Male), % | 17008(47.69) | 11849(48.74) | 0.04 |
| Ethnicity |  |  | < 0.0001 |
| Mexican American, % | 7035(9.85) | 3276(5.58) |  |
| Non-Hispanic Black, % | 6548( 9.55) | 6272(13.23) |  |
| Non-Hispanic White, % | 14757(66.41) | 10768(70.14) |  |
| Other Hispanic, % | 3258(6.52) | 1856(4.70) |  |
| Other Race, % | 4079(7.66) | 2018(6.35) |  |
| Age, year | 41.02±0.17 | 57.18±0.19 | < 0.0001 |
| WBC, 1000 cells/μL | 7.20±0.02 | 7.46±0.03 | < 0.0001 |
| LymP, % | 30.54±0.08 | 29.11±0.11 | < 0.0001 |
| MonP, % | 7.89±0.02 | 8.16±0.03 | < 0.0001 |
| SegneP, % | 58.18±0.09 | 59.13±0.11 | < 0.0001 |
| EoP, % | 2.73±0.01 | 2.92±0.02 | < 0.0001 |
| BaP, % | 0.71±0.00 | 0.74±0.01 | < 0.0001 |
| SeneP, % | 4.27±0.02 | 4.46±0.02 | < 0.0001 |
| Eo, 1000 cells/μL | 0.19±0.00 | 0.21±0.00 | < 0.0001 |
| RBC, million cells/μL | 4.72±0.01 | 4.68±0.01 | < 0.0001 |
| Hg, g/dl | 14.31±0.02 | 14.23±0.02 | < 0.0001 |
| Hem, % | 42.13±0.05 | 42.00±0.07 | 0.004 |
| MCV, fL | 89.46±0.06 | 89.90±0.08 | < 0.0001 |
| MCH, pg | 30.39±0.03 | 30.47±0.04 | 0.001 |
| MCHC, g/cL | 0.34±0.00 | 0.34±0.00 | < 0.0001 |
| RDW, % | 12.97±0.01 | 13.32±0.02 | < 0.0001 |
| Plt, 1000 cells/μL | 253.92±0.65 | 251.31±0.84 | 0.001 |
| MPV, fL | 8.18±0.01 | 8.21±0.01 | 0.005 |
| ALT, U/L | 24.54±0.15 | 26.15±0.18 | < 0.0001 |
| AST, U/L | 24.15±0.10 | 25.87±0.14 | < 0.0001 |
| Ca, mg/dL | 9.40±0.01 | 9.44±0.01 | < 0.0001 |
| TC, mg/dL | 193.50±0.38 | 197.66±0.52 | < 0.0001 |
| HCO<sub>3</sub>, mmol/L | 24.76±0.05 | 25.07±0.05 | < 0.0001 |
| GGT, U/L | 25.21±0.19 | 34.44±0.41 | < 0.0001 |
| TP, g/dL | 7.16±0.01 | 7.15±0.01 | 0.004 |
| TG, mg/dL | 136.74±1.03 | 170.49±1.39 | < 0.0001 |
| UA, mg/dL | 5.18±0.01 | 5.77±0.01 | < 0.0001 |
| Na, mmol/L | 139.36±0.06 | 139.36±0.07 | 0.86 |
| Lym, 1000 cells/μL | 2.14±0.01 | 2.15±0.02 | 0.69 |

HSW’s quartiles (HSWQ) were determined. The HSW were classified into Q1 [1.88,3.64] cL/g, Q2 (3.64,3.84] cL/g, Q3 (3.84,4.11] cL/g, and Q4 (4.11,11.74] cL/g. Details of the HSW quartiles were shown in **Table 2**. Hypertension prevalence in HSW quartiles was 28.59%, 33.35%, 39.37%, and 47.74%, respectively. The percentage of Hypertension increases with increasing HSW levels. And increasing HSW levels accompany decreasing males percentage and increased age. The general characteristics of sex and age were summarized. Males (36.63%) had a higher Hypertension prevalence (**Table S1**) than females (35.66%) with *P* <0.05. HSW level in females (3.92±0.01) is significantly higher than in males (3.82±0.00). Participants > 60 years had a higher (**Table S2**) hypertensive prevalence (67.64%) and HSW (3.99±0.01) than <= 60.

**Table 2.** General characteristics for quartiles of HSW.

|  | **Q1 (n=15,008)** | **Q2 (n=14,964)** | **Q3 (n=14,930)** | **Q4 (n=14,965)** | **P-value** |
| --- | --- | --- | --- | --- | --- |
| **HSW range, cL/g** | **[1.88,3.64]** | **(3.64,3.84]** | **(3.84,4.11]** | **(4.11,11.74]** |  |
| Hypertension, % | 4557(28.59) | 5476(33.35) | 6364(39.37) | 7793(47.74) | < 0.0001 |
| HSW, cL/g | 3.49±0.00 | 3.74±0.00 | 3.96±0.00 | 4.53±0.01 | < 0.0001 |
| COPD, % | 414(3.08) | 439(3.31) | 511(4.13) | 686(6.20) | < 0.0001 |
| DM state |  |  |  |  | < 0.0001 |
| Health, % | 10087(71.01) | 8536(61.23) | 6907(50.62) | 5399(41.03) |  |
| preDM, % | 3281(20.92) | 4320(27.98) | 5262(34.69) | 5631(36.90) |  |
| DM, % | 1637( 8.07) | 2104(10.79) | 2759(14.70) | 3928(22.08) |  |
| Anemia |  |  |  |  | < 0.0001 |
| Non-Anaemia | 14678(98.62) | 14404(97.56) | 13991(95.58) | 11109(77.62) |  |
| Mild | 306( 1.31) | 505( 2.20) | 818( 3.89) | 2534(14.42) |  |
| Moderate | 23(0.06) | 55(0.24) | 121(0.53) | 1243(7.55) |  |
| Severe | 1(0.00) | 0(0.00) | 0(0.00) | 79(0.41) |  |
| Sex, (Male), % | 7644(51.06) | 7765(52.02) | 7453(48.83) | 5995(37.02) | < 0.0001 |
| Ethnicity |  |  |  |  | < 0.0001 |
| Mexican American, % | 3203(8.25) | 2836(8.53) | 2397(8.60) | 1875(7.75) |  |
| Non-Hispanic Black, % | 1449( 4.54) | 2398( 8.12) | 3341(11.90) | 5632(23.30) |  |
| Non-Hispanic White, % | 8026(75.66) | 6976(71.15) | 5957(65.19) | 4566(53.91) |  |
| Other Hispanic, % | 1129(5.12) | 1231(5.59) | 1487(6.61) | 1267(6.50) |  |
| Other Race, % | 1201(6.43) | 1523(6.60) | 1748(7.71) | 1625(8.53) |  |
| Age, year | 42.20±0.26 | 45.89±0.24 | 49.61±0.28 | 52.09±0.29 | < 0.0001 |
| WBC, 1000 cells/μL | 7.14±0.03 | 7.21±0.03 | 7.36±0.04 | 7.59±0.04 | < 0.0001 |
| LymP, % | 30.28±0.09 | 30.33±0.10 | 29.97±0.11 | 29.27±0.15 | < 0.0001 |
| MonP, % | 7.86±0.03 | 7.99±0.03 | 8.06±0.03 | 8.12±0.03 | < 0.0001 |
| SegneP, % | 58.45±0.11 | 58.23±0.11 | 58.48±0.12 | 59.12±0.16 | < 0.0001 |
| EoP, % | 2.78±0.02 | 2.81±0.02 | 2.83±0.02 | 2.79±0.02 | 0.35 |
| BaP, % | 0.67±0.01 | 0.71±0.01 | 0.75±0.01 | 0.78±0.01 | < 0.0001 |
| SeneP, % | 4.24±0.02 | 4.27±0.02 | 4.38±0.03 | 4.54±0.03 | < 0.0001 |
| Eo, 1000 cells/μL | 0.20±0.00 | 0.20±0.00 | 0.20±0.00 | 0.21±0.00 | < 0.001 |
| RBC, million cells/μL | 4.68±0.01 | 4.73±0.01 | 4.73±0.01 | 4.70±0.01 | < 0.0001 |
| Hg, g/dl | 14.69±0.03 | 14.53±0.02 | 14.30±0.02 | 13.28±0.02 | < 0.0001 |
| Hem, % | 42.48±0.08 | 42.68±0.06 | 42.44±0.07 | 40.19±0.08 | < 0.0001 |
| MCV, fL | 91.03±0.08 | 90.48±0.06 | 89.86±0.08 | 85.95±0.13 | < 0.0001 |
| MCH, pg | 31.49±0.03 | 30.80±0.03 | 30.27±0.03 | 28.40±0.05 | < 0.0001 |
| MCHC, g/cL | 3.46±0.00 | 3.40±0.00 | 3.37±0.00 | 3.30±0.00 | < 0.0001 |
| RDW, % | 12.07±0.01 | 12.72±0.01 | 13.35±0.01 | 14.92±0.02 | < 0.0001 |
| Plt, 1000 cells/μL | 254.92±0.87 | 250.36±0.82 | 248.05±0.86 | 259.58±1.07 | < 0.0001 |
| MPV, fL | 8.06±0.02 | 8.17±0.01 | 8.25±0.01 | 8.34±0.02 | < 0.0001 |
| ALT, U/L | 26.58±0.17 | 25.82±0.29 | 24.78±0.22 | 22.25±0.21 | < 0.0001 |
| AST, U/L | 25.44±0.14 | 24.84±0.15 | 24.52±0.17 | 23.92±0.17 | < 0.0001 |
| Ca, mg/dL | 9.47±0.01 | 9.43±0.01 | 9.39±0.01 | 9.32±0.01 | < 0.0001 |
| TC, mg/dL | 196.81±0.51 | 197.00±0.58 | 194.79±0.52 | 189.62±0.73 | < 0.0001 |
| HCO3, mmol/L | 24.70±0.05 | 24.93±0.06 | 25.00±0.06 | 24.88±0.06 | < 0.0001 |
| GGT, U/L | 28.09±0.35 | 27.62±0.32 | 29.31±0.56 | 29.52±0.42 | < 0.001 |
| TP, g/dL | 7.20±0.01 | 7.17±0.01 | 7.12±0.01 | 7.12±0.01 | < 0.0001 |
| TG, mg/dL | 158.19±1.78 | 147.04±1.45 | 146.84±1.55 | 139.23±1.43 | < 0.0001 |
| UA, mg/dL | 5.36±0.02 | 5.40±0.01 | 5.41±0.02 | 5.41±0.02 | 0.06 |
| Na, mmol/L | 139.09±0.05 | 139.35±0.06 | 139.56±0.09 | 139.56±0.10 | < 0.0001 |
| Lym, 1000 cells/μL | 2.11±0.01 | 2.13±0.01 | 2.16±0.01 | 2.21±0.03 | 0.001 |

### 3.2 Logistic regression for HSW and Hypertension

The univariate Logistic details are shown in **Table S3.** And the adjusted HSW, HSWQ, RDW, and MCHC were shown in **Table 3**. Three models were utilized in the adjustment. The ORs value of HSW in the three multivariant adjusted models were still significant, including 1.34 (95%CI 1.25, 1.43), 1.1 6(95%CI 1.07, 1.25), and 1.23 (95%CI 1.11, 1.36). Taking Q1 as a reference and adjusting with multivariable (**Table 3**), Q4 was still significant and had the highest ORs, 1.28 (95% CI 1.13,1.45). In summary, higher HSW might be a risk of Hypertension and might promote the prevalence of Hypertension.

**Table 3.** The univariate and adjusted Logistic congression model among HSW, HSWQ, RDW, and MCHC.

| Variables | Hypertension |  |  |  |  |  |  |  |
| --- | --- | --- | --- | --- | --- | --- | --- | --- |
|  | OR (95% CI)<sup>a</sup> | P_value | OR (95% CI)<sup>b</sup> | P_value | OR (95% CI)<sup>c</sup> | P_value | OR (95% CI)<sup>d</sup> | P_value |
| HSW | 1.78(1.67,1.89) | <0.0001 | 1.34(1.25,1.43) | <0.0001 | 1.16(1.07,1.25) | <0.001 | 1.23(1.11,1.36) | <0.0001 |
| **HSWQ** |  |  |  |  |  |  |  |  |
| Q1 | ref | ref | ref | ref | ref | ref | ref | ref |
| Q2 | 1.25(1.17,1.33) | <0.0001 | 1.01(0.94,1.08) | 0.82 | 0.98(0.92,1.05) | 0.61 | 1.05(0.97,1.13) | 0.24 |
| Q3 | 1.62(1.51,1.74) | <0.0001 | 1.09(1.00,1.18) | 0.05 | 1.02(0.94,1.12) | 0.63 | 1.11(1.00,1.23) | 0.05 |
| Q4 | 2.28(2.13,2.44) | <0.0001 | 1.45(1.34,1.58) | <0.0001 | 1.19(1.07,1.32) | 0.001 | 1.28(1.13,1.45) | <0.001 |
| RDW | 1.25(1.22,1.28) | <0.0001 | 1.12(1.10,1.15) | <0.0001 | 1.07(1.05,1.10) | <0.0001 | 1.07(1.04,1.10) | <0.0001 |
| MCHC | 0.42(0.32,0.57) | <0.0001 | 0.75(0.54,1.02) | 0.07 | 1.70(1.18,2.43) | 0.004 | 5.25(0.67,41.13) | 0.11 |

a Model 1: unadjusted; b Model 2: adjusted with sex and age; c Model 3: adjusted with sex, age, ethnicity, COPD, DM state, and Anemia; d Model 4: adjusted with sex, age, ethnicity, COPD, DM state, Anemia, WBC, LymP, MonP, SegneP, EoP, BaP, SeneP, Eo, RBC, Hg, Hem, MCV, MCH, Plt, MPV, ALT, AST, Ca, TC, HCO<sub>3</sub>, GGT, TP, TG, and UA.

More importantly, The OR of RDW was consistently smaller than HSW’s, both univariate or adjusted. For example, the adjusted ORs with Model 4 in HSW and RDW were 1.23 (95% CI1.11,1.36) and 1.07 (95% CI 1.04,1.10). In conclusion, higher HSW is a risk for the evolution of Hypertension with a much higher predictive value than RDW.

### 3.3 The Cox regression

Among the 24,190 Hypertension patients, 3,184 were excluded because of the miss the following information. 5,487 Hypertension patients died, and 15,519 was alive during the follow-up time. Follow-up time ranged from 1 to 249 months, and the median and interquartile spacing of follow-up times were 97 (51, 151) months. The general information on dead and alive Hypertension patients was shown in **Table 4**. The HSW level in the dead Hypertension groups (3.99±0.01) is higher than in the alive Hypertension group (3.90±0.01). And the dead Hypertension group had a higher age, but the sex composition was insignificant.

**Table 4.** The general information on dead or alive Hypertension patients.

| Variables | Hypertensive patients alive (n=15,519) | Hypertensive patients dead (n=5,487) | P_value |
| --- | --- | --- | --- |
| HSW, cL/g | 3.90±0.01 | 3.99±0.01 | < 0.0001 |
| COPD, % | 760( 5.05) | 563(11.19) | < 0.0001 |
| DM state |  |  | < 0.0001 |
| Health, % | 5546(41.63) | 1793(35.21) |  |
| preDM, % | 5730(36.58) | 1696(31.13) |  |
| DM, % | 4241(21.79) | 1991(33.66) |  |
| Anemia |  |  | < 0.0001 |
| Non-Anaemia | 13917(93.06) | 4458(84.44) |  |
| Mild | 1228( 5.27) | 778(11.90) |  |
| Moderate | 353(1.60) | 246(3.60) |  |
| Severe | 21(0.07) | 5(0.07) |  |
| Sex, (Male), % | 7396(48.91) | 2894(47.68) | 0.13 |
| Ethnicity |  |  | < 0.0001 |
| Mexican American, % | 2340(6.24) | 680(3.30) |  |
| Non-Hispanic Black, % | 4087(13.56) | 1153(11.63) |  |
| Non-Hispanic White, % | 6355(68.60) | 3258(78.55) |  |
| Other Hispanic, % | 1325(4.89) | 231(3.02) |  |
| Other Race, % | 1412(6.71) | 165(3.49) |  |
| Age, year | 53.87±0.20 | 69.17±0.28 | < 0.0001 |
| WBC, 1000 cells/μL | 7.42±0.03 | 7.54±0.05 | 0.03 |
| LymP, % | 29.77±0.13 | 26.66±0.17 | < 0.0001 |
| MonP, % | 8.06±0.03 | 8.47±0.05 | < 0.0001 |
| SegneP, % | 58.60±0.13 | 61.22±0.17 | < 0.0001 |
| EoP, % | 2.90±0.02 | 3.00±0.04 | 0.06 |
| BaP, % | 0.74±0.01 | 0.69±0.01 | < 0.0001 |
| SeneP, % | 4.41±0.02 | 4.64±0.03 | < 0.0001 |
| Eo, 1000 cells/μL | 0.21±0.00 | 0.22±0.00 | 0.02 |
| RBC, million cells/μL | 4.72±0.01 | 4.52±0.01 | < 0.0001 |
| Hg, g/dl | 14.32±0.03 | 13.95±0.04 | < 0.0001 |
| Hem, % | 42.20±0.07 | 41.23±0.11 | < 0.0001 |
| MCV, fL | 89.58±0.09 | 91.42±0.11 | < 0.0001 |
| MCH, pg | 30.39±0.04 | 30.94±0.05 | < 0.0001 |
| MCHC, g/cL | 3.39±0.00 | 3.38±0.00 | 0.002 |
| RDW, % | 13.21±0.02 | 13.46±0.03 | < 0.0001 |
| Plt, 1000 cells/μL | 253.47±0.98 | 246.87±1.56 | < 0.0001 |
| MPV, fL | 8.21±0.02 | 8.17±0.02 | 0.03 |
| ALT, U/L | 27.21±0.21 | 23.44±0.39 | < 0.0001 |
| AST, U/L | 26.08±0.15 | 26.80±0.40 | 0.09 |
| Ca, mg/dL | 9.44±0.01 | 9.46±0.01 | 0.09 |
| TC, mg/dL | 199.25±0.60 | 197.62±0.81 | 0.07 |
| HCO3, mmol/L | 25.03±0.05 | 24.91±0.08 | 0.11 |
| GGT, U/L | 33.24±0.43 | 38.78±1.51 | < 0.001 |
| TP, g/dL | 7.15±0.01 | 7.17±0.01 | 0.04 |
| TG, mg/dL | 172.82±1.72 | 167.12±2.20 | 0.03 |
| UA, mg/dL | 5.73±0.02 | 5.97±0.03 | < 0.0001 |
| Na, mmol/L | 139.30±0.08 | 139.05±0.07 | 0.01 |
| Lym, 1000 cells/μL | 2.18±0.02 | 2.01±0.03 | < 0.0001 |

Other information among the quartiles of HSW among these Hypertension patients is shown in **Table 5**. The mortality rates of the Q1, Q2, Q3, and Q4 were 17.66%, 20.46%, 20.78%, and 25.02% (*P*<0.001). Mortality fluctuates as the quantile of HSW increases. With increasing HSW quartiles, the percentage of males decreased, and age increased. The mortality rate in sex was insignificant (**Table S4**). Hypertension patients (**Table S5**) with age > 60 had a higher mortality rate (36.75%) than those <= 60 (8.61%).

**Table 5.** The general information of HSW quartiles among dead or alive Hypertension patients.

| **Variables** | **Q1 (n=4,446)** | **Q2 (n=5,061)** | **Q3 (n=5,372)** | **Q4 (n=6,127)** | **P-value** |
| --- | --- | --- | --- | --- | --- |
| **HSW range, cL/g** | **[1.88,3.64]** | **(3.64,3.84]** | **(3.84,4.11]** | **(4.11,11.74]** |  |
| Mortality rate, % | 1018(17.66) | 1348(20.46) | 1409(20.78) | 1712(25.02) | < 0.0001 |
| HSW, cL/g | 3.50±0.00 | 3.74±0.00 | 3.96±0.00 | 4.53±0.01 | < 0.0001 |
| COPD, % | 227(5.38) | 258(5.04) | 334(6.15) | 504(8.95) | < 0.0001 |
| DM state |  |  |  |  | < 0.0001 |
| Health, % | 2098(52.61) | 2007(44.67) | 1729(35.49) | 1505(27.38) |  |
| preDM, % | 1361(30.55) | 1753(34.48) | 2034(38.66) | 2278(38.36) |  |
| DM, % | 986(16.84) | 1299(20.85) | 1608(25.85) | 2339(34.26) |  |
| Anemia |  |  |  |  | < 0.0001 |
| Non-Anaemia | 4280(97.84) | 4752(95.96) | 4884(93.54) | 4459(76.78) |  |
| Mild | 149( 1.99) | 274( 3.54) | 416( 5.52) | 1167(16.18) |  |
| Moderate | 17(0.17) | 35(0.49) | 72(0.94) | 475(6.75) |  |
| Severe | 0(0.00) | 0(0.00) | 0(0.00) | 26(0.28) |  |
| Sex, (Male), % | 2287(53.08) | 2574(50.46) | 2698(49.63) | 2731(40.95) | < 0.0001 |
| Ethnicity |  |  |  |  | < 0.0001 |
| Mexican American, % | 800(5.66) | 843(5.92) | 718(5.50) | 659(5.41) |  |
| Non-Hispanic Black, % | 452( 4.61) | 919( 9.38) | 1367(13.73) | 2502(25.79) |  |
| Non-Hispanic White, % | 2589(79.14) | 2586(75.38) | 2397(70.10) | 2041(57.17) |  |
| Other Hispanic, % | 307(4.29) | 348(4.16) | 465(5.08) | 436(4.47) |  |
| Other Race, % | 298(6.30) | 365(5.15) | 425(5.60) | 489(7.15) |  |
| Age, year | 52.83±0.34 | 56.51±0.33 | 58.75±0.30 | 60.47±0.33 | < 0.0001 |
| WBC, 1000 cells/μL | 7.25±0.04 | 7.24±0.04 | 7.58±0.05 | 7.74±0.07 | < 0.0001 |
| LymP, % | 29.66±0.17 | 29.47±0.15 | 29.01±0.20 | 28.28±0.22 | < 0.0001 |
| MonP, % | 8.03±0.05 | 8.14±0.06 | 8.17±0.04 | 8.24±0.05 | 0.01 |
| SegneP, % | 58.76±0.19 | 58.78±0.16 | 59.24±0.23 | 59.86±0.23 | < 0.001 |
| EoP, % | 2.92±0.03 | 2.94±0.04 | 2.92±0.04 | 2.92±0.03 | 0.96 |
| BaP, % | 0.68±0.01 | 0.72±0.01 | 0.74±0.01 | 0.78±0.01 | < 0.0001 |
| SeneP, % | 4.31±0.03 | 4.31±0.03 | 4.55±0.04 | 4.67±0.04 | < 0.0001 |
| Eo, 1000 cells/μL | 0.21±0.00 | 0.21±0.00 | 0.22±0.00 | 0.22±0.00 | 0.01 |
| RBC, million cells/μL | 4.67±0.01 | 4.69±0.01 | 4.70±0.01 | 4.67±0.01 | 0.01 |
| Hg, g/dl | 14.76±0.04 | 14.50±0.03 | 14.29±0.03 | 13.36±0.03 | < 0.0001 |
| Hem, % | 42.61±0.11 | 42.55±0.08 | 42.33±0.09 | 40.39±0.10 | < 0.0001 |
| MCV, fL | 91.50±0.10 | 90.97±0.09 | 90.24±0.11 | 86.95±0.17 | < 0.0001 |
| MCH, pg | 31.71±0.04 | 30.99±0.04 | 30.46±0.04 | 28.76±0.06 | < 0.0001 |
| MCHC, g/cL | 3.47±0.00 | 3.41±0.00 | 3.38±0.00 | 3.31±0.00 | < 0.0001 |
| RDW, % | 12.11±0.01 | 12.73±0.01 | 13.38±0.01 | 14.94±0.03 | < 0.0001 |
| Plt, 1000 cells/μL | 255.60±1.46 | 250.77±1.34 | 247.13±1.42 | 254.88±1.63 | < 0.0001 |
| MPV, fL | 8.06±0.02 | 8.17±0.02 | 8.27±0.02 | 8.32±0.02 | < 0.0001 |
| ALT, U/L | 29.23±0.33 | 26.84±0.36 | 26.30±0.52 | 23.06±0.25 | < 0.0001 |
| AST, U/L | 27.37±0.27 | 26.02±0.24 | 26.29±0.38 | 25.12±0.29 | < 0.0001 |
| Ca, mg/dL | 9.50±0.01 | 9.47±0.01 | 9.44±0.01 | 9.38±0.01 | < 0.0001 |
| TC, mg/dL | 204.54±0.84 | 201.19±0.89 | 197.88±0.83 | 191.40±1.02 | < 0.0001 |
| HCO3, mmol/L | 24.87±0.07 | 25.02±0.07 | 25.04±0.08 | 25.10±0.08 | 0.08 |
| GGT, U/L | 36.37±0.83 | 32.65±0.65 | 34.53±1.35 | 33.95±0.73 | 0.01 |
| TP, g/dL | 7.19±0.01 | 7.16±0.01 | 7.13±0.01 | 7.12±0.01 | < 0.0001 |
| TG, mg/dL | 191.31±3.51 | 169.47±2.27 | 168.14±2.65 | 156.15±2.51 | < 0.0001 |
| UA, mg/dL | 5.79±0.03 | 5.74±0.03 | 5.75±0.03 | 5.83±0.03 | 0.16 |
| Na, mmol/L | 139.00±0.07 | 139.17±0.07 | 139.39±0.11 | 139.45±0.12 | 0.004 |
| Lym, 1000 cells/μL | 2.11±0.01 | 2.10±0.02 | 2.16±0.02 | 2.19±0.05 | 0.04 |

The univariate Cox regression results was shown in **Table S6.** The details of HSW, HSWQ, RDW, and MCHC were shown in **Table 6**. Adjusted with multivariable, the HR value of HSW was still significant, 1.91(95% CI 1.69,2.16). Using Q1 as a reference and adjusted with multivariable (**Table 6**), Q2, Q3, and Q4 still had significant HRs, 1.16 (95% CI 1.04,1.29), 1.46 (95% CI 1.31,1.63), and 2.35 (95% CI 2.06,2.69), respectively. The higher the HSW quartile was, the poor the Hypertension participants’ prognosis might be.

**Table 6.** The univariate and adjusted Cox congression model among HSW, HSWQ, RDW, and MCHC.

| Variables | Hypertension mortality |  |  |  |  |  |  |  |
| --- | --- | --- | --- | --- | --- | --- | --- | --- |
|  | HR (95% CI)<sup>a</sup> | P_value | HR (95% CI)<sup>b</sup> | P_value | HR (95% CI)<sup>c</sup> | P_value | HR (95% CI)<sup>d</sup> | P_value |
| HSW | 2.02(1.88,2.17) | <0.0001 | 1.91(1.75,2.07) | <0.0001 | 1.76(1.60,1.95) | <0.0001 | 1.91(1.69,2.16) | <0.0001 |
| **HSWQ** |  |  |  |  |  |  |  |  |
| Q1 | ref | ref | ref | ref | ref | ref | ref | ref |
| Q2 | 1.40(1.26,1.55) | <0.0001 | 1.09(0.98,1.21) | 0.11 | 1.09(0.98,1.22) | 0.13 | 1.16(1.04,1.29) | 0.01 |
| Q3 | 2.08(1.88,2.31) | <0.0001 | 1.45(1.31,1.60) | <0.0001 | 1.41(1.27,1.57) | <0.0001 | 1.46(1.31,1.63) | <0.0001 |
| Q4 | 3.31(2.96,3.71) | <0.0001 | 2.33(2.10,2.59) | <0.0001 | 2.08(1.84,2.34) | <0.0001 | 2.35(2.06,2.69) | <0.0001 |
| RDW | 1.29(1.26,1.32) | <0.0001 | 1.26(1.22,1.29) | <0.0001 | 1.22(1.18,1.26) | <0.0001 | 1.22(1.18,1.27) | <0.0001 |
| MCHC | 0.16(0.10,0.25) | <0.0001 | 0.31(0.20,0.47) | <0.0001 | 0.53(0.34,0.82) | 0.004 | 0.60(0.02,22.64) | 0.78 |

a Model 1: unadjusted; b Model 2: adjusted with sex and age; c Model 3: adjusted with sex, age, ethnicity, COPD, DM state, and Anemia; d Model 4: adjusted with sex, age, ethnicity, COPD, DM state, Anemia, WBC, LymP, MonP, SegneP, EoP, BaP, SeneP, Eo, RBC, Hg, Hem, MCV, MCH, Plt, MPV, ALT, AST, Ca, TC, HCO<sub>3</sub>, GGT, TP, TG, and UA.

More importantly, The HR of RDW was consistently smaller than HSW’s, both univariate or adjusted. For example, the adjusted HRs with the same multivariable in HSW and RDW were 1.91 (95% CI 1.69,2.16) and 1.22 (95% CI 1.18,1.27). In conclusion, higher HSW is a risk for the prognosis of Hypertension, accompanying a much higher predictive value than RDW.

### 3.4 Cox regression for HSW and Hypertension with subgroup

#### 3.4.1 Prognosis of HSW in Hypertensive patients after Sex and Age grouping

As shown in **Table S7** and **Table S8,** the mortality of Hypertension is significantly related to the stratification of age and sex. When adjusted with multivariate, HSW was still considerably correlated with Hypertension in males (HR: 1.53; 95% CI 1.24, 1.89) and females (HR: 1.48; 95% CI 1.17, 1.87). The HRs values of males were higher than that of females, indicating Hypertension male patients might be a poorer prognosis than females with an increasing HSW level.

In the age with multivariable adjustment, the HR of Hypertensive patients <= 60 years (HR: 2.25; 95% CI 1.78,2.85) was higher than >60 (HR: 1.37; 95% 1.15,1.63), indicating Hypertension patients <=60 years might be a poorer prognosis than >60 with an increasing HSW level.

#### 3.4.2 Prognosis of HSW in Hypertensive patients with different DM states

As shown in **Table S9,** three DM state was classified in this study, including healthy individuals, preDM, and DM patients. With adjustment with the same multivariate, the three DM states still showed significant HRs, including 1.60 (95 CI% 1.27,2.01), 1.69 (95% CI 1.23,2.33), and 1.36 (95% CI 1.13,1.64). Processing Q1 as a reference, Q4s of the three DM states were still significant, 1.90 (95% CI 1.44,2.50), 1.47 (95% CI1.03,2.08), and 1.57 (95% CI 1.22,2.02), respectively. In a word, when Hypertensive patients with the three DM states, a higher HSW predicted a poor prognosis.

### 3.5 RCS analysis and KM survival curve

In RCS analysis, according to the minimum fluctuation HR value range of 95% CI, the optimal value of HSW, MCHC, and RDW was determined (**Fig 3. A**). Both the RDW and HSW were poor prognoses of Hypertensive patients. The higher HSW and RDW accompany the increasing HRs (**Fig 3. A**). Inversely, higher MCHC accompany decreasing HRs. And the optimal inflexion point of HSW, MCHC, and RDW were 3.89, 3.37, and 13.01, respectively.

**Fig 3.**
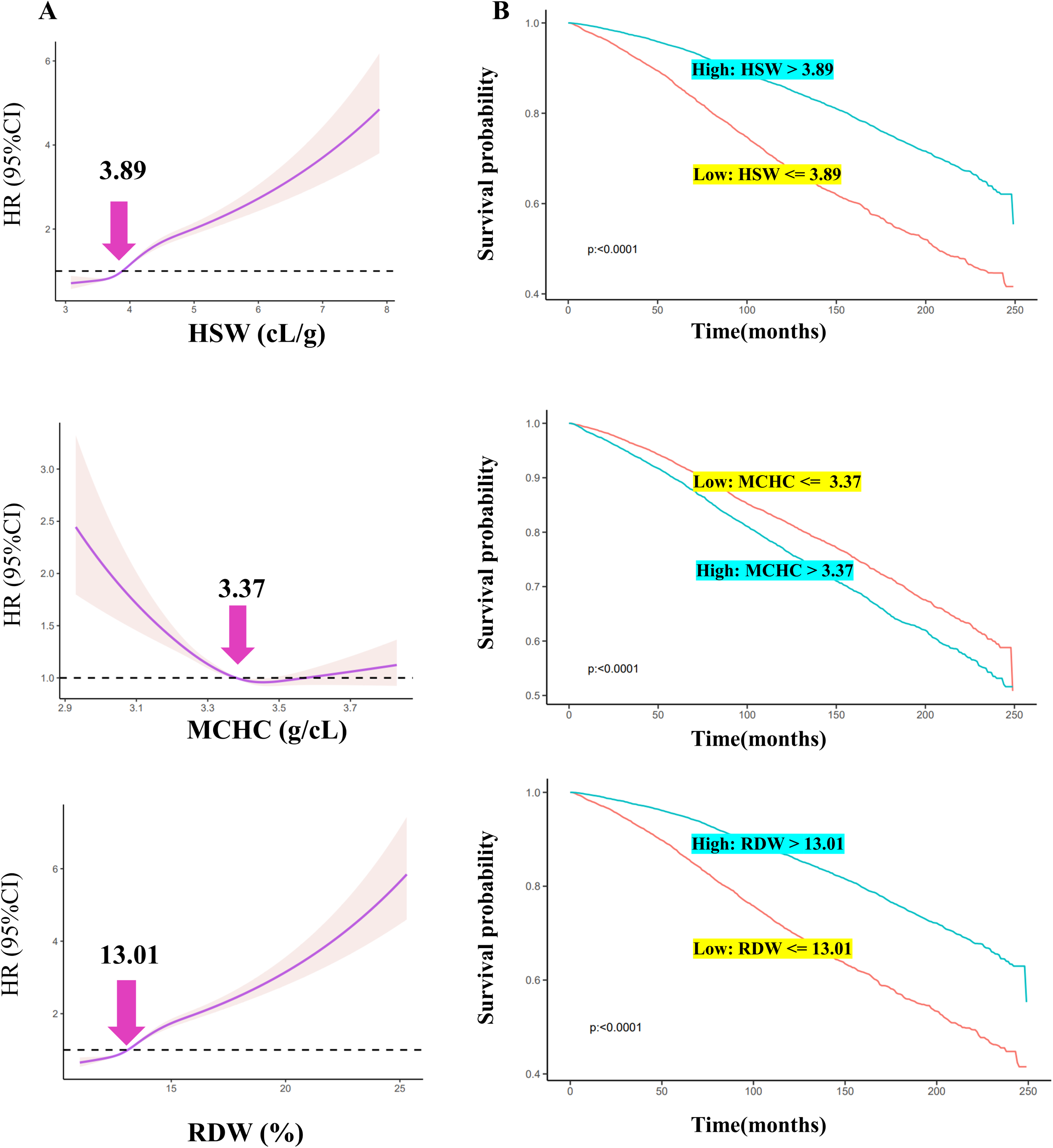
RCS analysis and KM survival curve. A The RCS for optimal cut-off HR value; B KM survival curve.

According to the optimal inflexion point, they were divided into the high-expression or low-expression groups. For example, an individual with an HSW >3.89 was classified into the high HSW group, else the low HSW group. Classified methods for RDW and MCHC were similar to HSW. Hypertensive patients with HSW > 3.89, MCHC <3.37, and RDW > 13.01 have poor survival rates (**Fig 3. B**). The three factors stated an excellent predictive value in the survival analysis of Hypertensive patients.

## 4. Discussion

This is the first article focusing on the clinical value of HSW with hypertension. It not only developed a calculation strategy for HSW but also verified the importance of HSW for the prevalence and long-term prognosis of hypertension. The prevalence of hypertension varied significantly among quartiles of HSW. And Higher levels of HSW account for a higher prevalence of hypertension. In hypertension patients, the risk of sex, age, and diabetes state stratification was different. Males had a higher risk of poor prognosis than females in hypertensive patients HSW increased. Individuals <=60 years old had a higher risk of poor prognosis than those >60 when HSW grew.

Furthermore, the prognosis of hypertension fluctuated significantly among quartiles of HSW and higher levels of HSW performance for a higher mortality rate of hypertension. Moreover, an inverse L-curve with an inflexion point of 3.89 was observed between HR and hypertension. And hypertensive patients with HSW > 3.89 indicated higher mortality and poor prognosis than HSW <= 3.89. These findings indicate that HSW can be used as a monitoring indicator of hypertensive incidence and prognosis.

Although HSW is calculated from RDW and MCHC, it has a much higher OR and HR value than both. The adjusted ORs and HR for MCHC were insignificant. But for the adjusted by Model 4 of HSW and RDW (**Table 3**), the ORs were 1.23 and 1.07. For HRs (**Table 6**), these were 1.91 and 1.22. In a word, HSW is not only an independent predictive factor of the prevalence and prognosis of hypertension, but the value is much higher than the RDW and MCHC.

The role of HSW in other diseases needs to be further investigated. For example, RDW ^27^ and MCHC ^28^ have already been recognized as diagnostic indicators of anaemia, and HSW is calculated from MCHC and RDW. MCHC mirrors the haemoglobin concentration and is related to an adverse poor prognosis in heart failure ^29^. Higher RDW ^2^ mirrors the potential deregulation of RBC balance, including weakened erythropoiesis and disorder RBC survival. Similarly, an increasing HSW might reflect a chaotic hematopoietic and oxygen-carrying. Because haemoglobin is an oxygen carrier, a rising HSW might mean an insufficient oxygen supply ^30^, which could trigger bad results ^31,32^. Therefore, HSW might have a specific value in anaemia and hypoxic sensitivity diseases, and more studies are needed for in-depth analysis.

RDW has received an increasing focus on cardiovascular disease but few on hypertension over the last decade. Higher RDW might result in thrombotic disorders ^33^ and poor complications in cardiovascular diseases, such as stroke ^34^ and acute myocardial infarction ^34^. Similarly, a prospective study from Turkey with 1202 newly hypertensive patients indicated RDW is a higher risk for major adverse cardiac events ^35^. Dispponntantly, this work did not optimize the cut-off value of RDW. Not only in cardio events the RDW can predict but also in early-stage renal function damage^36^. A cross-sectional study with 495 individuals ^37^ emphasized that a combination of high RDW and thrombophilia abnormalities had a risk of cerebral vein thrombosis with 33.20 ORs. In a 15-year follow-up retrospective cohort with more than twenty thousand adult participants ^38^ without a heart attack, stroke, or heart failure, increasing RDW was related to long-term heart failure incidence with an HR value of 1.47. The erythrocyte membrane enriches abundant cholesterol ^39^. Supplementing RBC within the atherosclerotic plaque will deteriorate plaque evolution ^40^, resulting in cardiovascular disease events. In summary, few studies contributed to the association of RDW among hypertensive patients. Further research could focus on this potential.

Another RDW-related novel index, RAR (the RDW-serum albumin ratio), has received increasing attention as an inflammation-related indicator in recent years. However, RAR was only utilized for the short-term prognosis in diseases (30 days ^41^, 90 days ^42,43^, 1 year ^44^). In hypertensive patients, HSW had over 97 months of long-term prognosis. Although RAR has some clinical value ^42,45^, they are combined with multiple tests. For example, the RDW ^42,45^ is derived from hematology, but serum albumin is from biochemistry, which may increase the patient’s financial burden. Unlike the above tests, both RDW and MCHC were included in hematology. In a word, HSW is more clinically actionable, applicable, and less financially burdensome for hypertensive patients.

This study has four strengths: innovation of HSW, large sample size, robust and credible results, and a combination of cross-sectional and prospective studies. Firstly, HSW is proposed by us for the first time in hypertensive patients, and the calculation strategy is formulated. Secondly, this study’s sample size and duration time (1999-2020) are large enough. NHANES applied a circle years weight to represent the American population. Thirdly, 30 covariates were included for adjustment to ensure the robustness and reliability of the results. More importantly, NHANES employed professional and trained personnel with standardized procedures to reduce deviation, such as strict physical examination and standard questionnaires. Finally, the combination of cross-sectional and prospective studies focuses on the incidence and prognosis between HSW and hypertensive patients. The optimal cut-off value for HSW was determined in prognosis analysis, and its long-term prognostic value was analyzed.

Furthermore, two limitations should be noted. First, the causality could not be obtained, and the current result should be approached carefully. Further prospective work is indispensable to verify the specific cause between HSW and hypertensive patients. Although many covariates were controlled and adjusted, there may still be potential and neglected confounders, like dietary patterns. Then, the HSW was only detected and calculated at a single point ^46^, which may trigger the misclassification. Nevertheless, this misclassification would lead to an underestimate but not an overestimation ^47^.

## 5. Conclusion

Higher HSW may be more likely to develop hypertension. And in hypertensive patients, higher HSW indicated a poor prognosis. The HSW predictive value is much higher than RDW and MCHC. Individuals male and aged <= 60 years might have a poor prognosis of hypertensive patients when HSW increases. In conclusion, HSW is an innovative independent risk factor for hypertensive prevalence and long-term prognosis.

## Abbreviations

ALT: alanine transaminase
BaP: basophils percentage
Ca: total calcium
COPD: chronic obstructive pulmonary disease
DM: diabetes mellitus
Eo: eosinophils number count
EoP: eosinophils percentage
EV-SD: erythrocyte volume’s standard deviation
GGT: gammaglutamyl transferase
Hb: hemoglobin
HCO3: bicarbonate
Hem: hematocrit
HR: hazard ratios
HSW: Hemoglobin specific volume width
HSWQ: HSW’s quartiles
KM: Kaplan–Meier
MCH: mean cell hemoglobin
MCHC: mean corpuscular haemoglobin concentration
MCQ: medical conditions questionnaire
MCV: mean corpuscular volume
MonP: monocyte percentage
MPV: mean platelet volume
Na: sodium
NHANES: national health and nutrition examination survey
OR: odds ratio
Plt: platelet count count
RBC: red blood cells
RCS: restricted cubic spline
RDW: red blood distribution width
SegneP: segmented neutrophils percentage.
TC: total cholesterol
TG: triglycerides
TP: total protein;
UA: uric acid.

## Declarations

### Ethics approval and consent to participate

The NHANES protocol was approved by the Ethics Review Committee of the National Center for Health Statistics.

### Consent for publication

All patients/participants includec in this study provided their written informed consent.

### Availability of data and materials

NHANES data were derived from https://www.cdc.gov/nchs/nhanes/index.htm. And the mortality data were linked to https://www.cdc.gov/nchs/ndi/index.htm.

### Author’s contributions

LZ and JC wrote the original draft. LZ, JC, KYW, YL, and LMW performed the research. LZ, YZ, ZFF, and LLZ analyzed the data. XMZ, ZFF, LFH, and XMG designed the experiment and revised the manuscript.

## Acknowledgements

Thanks to Lifeng Han for proposing the idea of the article and Xiumei Gao for optimizing the framework of the article. Thanks to the NHANES.

## References

1. Hernández Hernández JD, Villaseñor OR, Del Rio Alvarado J, Lucach RO, Zárate A, Saucedo R, Hernández-Valencia M. Morphological changes of red blood cells in peripheral blood smear of patients with pregnancy-related hypertensive disorders. Arch Med Res. 2015;46:479–483. doi: 10.1016/j.arcmed.2015.07.003

2. Salvagno GL, Sanchis-Gomar F, Picanza A, Lippi G. Red blood cell distribution width: A simple parameter with multiple clinical applications. Crit Rev Clin Lab Sci. 2015;52:86–105. doi: 10.3109/10408363.2014.992064

3. Pytel E, Duchnowicz P, Jackowska P, Wojdan K, Koter-Michalak M, Broncel M. Disorders of erythrocyte structure and function in hypertensive patients. Med Sci Monit. 2012;18:Br331–336. doi: 10.12659/msm.883265

4. Vokurková M, Nováková O, Dobesová Z, Kunes J, Zicha J. Relationships between membrane lipids and ion transport in red blood cells of Dahl rats. Life Sci. 2005;77:1452–1464. doi: 10.1016/j.lfs.2005.03.014

5. Lee SG, Rim JH, Kim JH. Association of hemoglobin levels with blood pressure and hypertension in a large population-based study: the Korea National Health and Nutrition Examination Surveys 2008-2011. Clin Chim Acta. 2015;438:12–18. doi: 10.1016/j.cca.2014.07.041

6. Kim NH, Lee JM, Kim HC, Lee JY, Yeom H, Lee JH, Suh I. Cross-sectional and longitudinal association between hemoglobin concentration and hypertension: A population-based cohort study. Medicine (Baltimore). 2016;95:e5041. doi: 10.1097/md.0000000000005041

7. Vega-Sánchez R, Tolentino-Dolores MC, Cerezo-Rodríguez B, Chehaibar-Besil G, Flores-Quijano ME. Erythropoiesis and Red Cell Indices Undergo Adjustments during Pregnancy in Response to Maternal Body Size but not Inflammation. Nutrients. 2020;12. doi: 10.3390/nu12040975

8. Seo SG, Lee MY, Park SH, Han JM, Lee KB, Kim H, Hyun YY. The association between red cell distribution width and incident hypertension in Korean adults. Hypertens Res. 2020;43:55–61. doi: 10.1038/s41440-019-0334-3

9. Van Craenenbroeck EM, Conraads VM, Greenlaw N, Gaudesius G, Mori C, Ponikowski P, Anker SD. The effect of intravenous ferric carboxymaltose on red cell distribution width: a subanalysis of the FAIR-HF study. Eur J Heart Fail. 2013;15:756–762. doi: 10.1093/eurjhf/hft068

10. Smeulders MJ, van den Berg S, Oudeman J, Nederveen AJ, Kreulen M, Maas M. Reliability of in vivo determination of forearm muscle volume using 3.0 T magnetic resonance imaging. J Magn Reson Imaging. 2010;31:1252–1255. doi: 10.1002/jmri.22153

11. Garcia-Hernandez A, Roldan-Cruz C, Vernon-Carter EJ, Alvarez-Ramirez J. Effects of leavening agent and time on bread texture and in vitro starch digestibility. J Food Sci Technol. 2022;59:1922–1930. doi: 10.1007/s13197-021-05206-1

12. Kibkalo I. Effectiveness of and Perspectives for the Sedimentation Analysis Method in Grain Quality Evaluation in Various Cereal Crops for Breeding Purposes. Plants (Basel). 2022;11. doi: 10.3390/plants11131640

13. Özkaya B, Baumgartner B, Özkaya H. Effects of concentrated and dephytinized wheat bran and rice bran addition on bread properties. J Texture Stud. 2018;49:84–93. doi: 10.1111/jtxs.12286

14. Liu Y, Zhang Q, Wang Y, Xu P, Wang L, Liu L, Rao Y. Enrichment of Wheat Bread with Platycodon grandiflorus Root (PGR) Flour: Rheological Properties and Microstructure of Dough and Physicochemical Characterization of Bread. Foods. 2023;12. doi: 10.3390/foods12030580

15. Zhang Y, Liu C, Yang M, Ou Z, Lin Y, Zhao F, Han S. Characterization and application of a novel xylanase from Halolactibacillus miurensis in wholewheat bread making. Front Bioeng Biotechnol. 2022;10:1018476. doi: 10.3389/fbioe.2022.1018476

16. Verdonck C, De Bondt Y, Pradal I, Bautil A, Langenaeken NA, Brijs K, Goos P, De Vuyst L, Courtin CM. Impact of process parameters on the specific volume of wholemeal wheat bread made using sourdough- and baker’s yeast-based leavening strategies. Int J Food Microbiol. 2023;396:110193. doi: 10.1016/j.ijfoodmicro.2023.110193

17. Ji X, Ke W. Red blood cell distribution width and all-cause mortality in congestive heart failure patients: a retrospective cohort study based on the Mimic-III database. Front Cardiovasc Med. 2023;10:1126718. doi: 10.3389/fcvm.2023.1126718

18. Danese E, Lippi G, Montagnana M. Red blood cell distribution width and cardiovascular diseases. J Thorac Dis. 2015;7:E402–411. doi: 10.3978/j.issn.2072-1439.2015.10.04

19. Chauhan K, Bisht B, Kathuria K, Bisht R, Hatwal V. Z score analysis: A novel approach to interpretation of an erythrogram. Indian J Pathol Microbiol. 2023;66:85–90. doi: 10.4103/ijpm.ijpm_1188_21

20. Gredic M, Blanco I, Kovacs G, Helyes Z, Ferdinandy P, Olschewski H, Barberà JA, Weissmann N. Pulmonary hypertension in chronic obstructive pulmonary disease. Br J Pharmacol. 2021;178:132–151. doi: 10.1111/bph.14979

21. Kovacs G, Avian A, Bachmaier G, Troester N, Tornyos A, Douschan P, Foris V, Sassmann T, Zeder K, Lindenmann J, et al. Severe Pulmonary Hypertension in COPD: Impact on Survival and Diagnostic Approach. Chest. 2022;162:202–212. doi: 10.1016/j.chest.2022.01.031

22. Ferrannini E, Cushman WC. Diabetes and hypertension: the bad companions. Lancet. 2012;380:601–610. doi: 10.1016/s0140-6736(12)60987-8

23. Li M, Bertout JA, Ratcliffe SJ, Eckenhoff MF, Simon MC, Floyd TF. Acute anemia elicits cognitive dysfunction and evidence of cerebral cellular hypoxia in older rats with systemic hypertension. Anesthesiology. 2010;113:845–858. doi: 10.1097/ALN.0b013e3181eaaef9

24. De Franceschi L, Iolascon A, Taher A, Cappellini MD. Clinical management of iron deficiency anemia in adults: Systemic review on advances in diagnosis and treatment. Eur J Intern Med. 2017;42:16–23. doi: 10.1016/j.ejim.2017.04.018

25. Feng M, McSparron JI, Kien DT, Stone DJ, Roberts DH, Schwartzstein RM, Vieillard-Baron A, Celi LA. Transthoracic echocardiography and mortality in sepsis: analysis of the MIMIC-III database. Intensive Care Med. 2018;44:884–892. doi: 10.1007/s00134-018-5208-7

26. Liu X, Zhang D, Liu Y, Sun X, Han C, Wang B, Ren Y, Zhou J, Zhao Y, Shi Y, et al. Dose-Response Association Between Physical Activity and Incident Hypertension: A Systematic Review and Meta-Analysis of Cohort Studies. Hypertension. 2017;69:813–820. doi: 10.1161/hypertensionaha.116.08994

27. Taneri PE, Gómez-Ochoa SA, Llanaj E, Raguindin PF, Rojas LZ, Roa-Díaz ZM, Salvador D, Jr., Groothof D, Minder B, Kopp-Heim D, et al. Anemia and iron metabolism in COVID-19: a systematic review and meta-analysis. Eur J Epidemiol. 2020;35:763–773. doi: 10.1007/s10654-020-00678-5

28. Haider BA, Olofin I, Wang M, Spiegelman D, Ezzati M, Fawzi WW. Anaemia, prenatal iron use, and risk of adverse pregnancy outcomes: systematic review and meta-analysis. Bmj. 2013;346:f3443. doi: 10.1136/bmj.f3443

29. Choy M, Zhen Z, Dong B, Chen C, Dong Y, Liu C, Liang W, Xue R. Mean corpuscular haemoglobin concentration and outcomes in heart failure with preserved ejection fraction. ESC Heart Fail. 2023;10:1214–1221. doi: 10.1002/ehf2.14225

30. Crespo-Leiro MG, Metra M, Lund LH, Milicic D, Costanzo MR, Filippatos G, Gustafsson F, Tsui S, Barge-Caballero E, De Jonge N, et al. Advanced heart failure: a position statement of the Heart Failure Association of the European Society of Cardiology. Eur J Heart Fail. 2018;20:1505–1535. doi: 10.1002/ejhf.1236

31. Grieco DL, Maggiore SM, Roca O, Spinelli E, Patel BK, Thille AW, Barbas CSV, de Acilu MG, Cutuli SL, Bongiovanni F, et al. Non-invasive ventilatory support and high-flow nasal oxygen as first-line treatment of acute hypoxemic respiratory failure and ARDS. Intensive Care Med. 2021;47:851–866. doi: 10.1007/s00134-021-06459-2

32. Punj S, Ghafourian K, Ardehali H. Iron deficiency and supplementation in heart failure and chronic kidney disease. Mol Aspects Med. 2020;75:100873. doi: 10.1016/j.mam.2020.100873

33. Montagnana M, Cervellin G, Meschi T, Lippi G. The role of red blood cell distribution width in cardiovascular and thrombotic disorders. Clin Chem Lab Med. 2011;50:635–641. doi: 10.1515/cclm.2011.831

34. Feng GH, Li HP, Li QL, Fu Y, Huang RB. Red blood cell distribution width and ischaemic stroke. Stroke Vasc Neurol. 2017;2:172–175. doi: 10.1136/svn-2017-000071

35. Uzun F, Güner A, Pusuroglu H, Demir AR, Gündüz S, Gürbak İ, Aslan S, Demirci G, Gültekin Güner E, Arslan E, et al. Association of red blood cell distribution width, systemic-immune-inflammation index and poor cardiovascular outcomes in patients with newly diagnosed hypertension. Clin Exp Hypertens. 2022;44:530–538. doi: 10.1080/10641963.2022.2079668

36. Li ZZ, Chen L, Yuan H, Zhou T, Kuang ZM. Relationship between red blood cell distribution width and early-stage renal function damage in patients with essential hypertension. J Hypertens. 2014;32:2450–2455; discussion 2456. doi: 10.1097/hjh.0000000000000356

37. Maino A, Abbattista M, Bucciarelli P, Artoni A, Passamonti SM, Lanfranconi S, Martinelli I. Red cell distribution width and the risk of cerebral vein thrombosis: A case-control study. Eur J Intern Med. 2017;38:46–51. doi: 10.1016/j.ejim.2016.10.017

38. Borné Y, Smith JG, Melander O, Hedblad B, Engström G. Red cell distribution width and risk for first hospitalization due to heart failure: a population-based cohort study. Eur J Heart Fail. 2011;13:1355–1361. doi: 10.1093/eurjhf/hfr127

39. Kolodgie FD, Burke AP, Nakazawa G, Cheng Q, Xu X, Virmani R. Free cholesterol in atherosclerotic plaques: where does it come from? Curr Opin Lipidol. 2007;18:500–507. doi: 10.1097/MOL.0b013e3282efa35b

40. Kolodgie FD, Gold HK, Burke AP, Fowler DR, Kruth HS, Weber DK, Farb A, Guerrero LJ, Hayase M, Kutys R, et al. Intraplaque hemorrhage and progression of coronary atheroma. N Engl J Med. 2003;349:2316–2325. doi: 10.1056/NEJMoa035655

41. Lu C, Long J, Liu H, Xie X, Xu D, Fang X, Zhu Y. Red blood cell distribution width-to-albumin ratio is associated with all-cause mortality in cancer patients. J Clin Lab Anal. 2022;36:e24423. doi: 10.1002/jcla.24423

42. Ni Q, Wang X, Wang J, Chen P. The red blood cell distribution width-albumin ratio: A promising predictor of mortality in heart failure patients - A cohort study. Clin Chim Acta. 2022;527:38–46. doi: 10.1016/j.cca.2021.12.027

43. Seo YJ, Yu J, Park JY, Lee N, Lee J, Park JH, Kim HY, Kong YG, Kim YK. Red cell distribution width/albumin ratio and 90-day mortality after burn surgery. Burns Trauma. 2022;10:tkab050. doi: 10.1093/burnst/tkab050

44. Weng Y, Peng Y, Xu Y, Wang L, Wu B, Xiang H, Ji K, Guan X. The Ratio of Red Blood Cell Distribution Width to Albumin Is Correlated With All-Cause Mortality of Patients After Percutaneous Coronary Intervention - A Retrospective Cohort Study. Front Cardiovasc Med. 2022;9:869816. doi: 10.3389/fcvm.2022.869816

45. Lin Z, Zhao Y, Xiao L, Qi C, Chen Q, Li Y. Blood urea nitrogen to serum albumin ratio as a new prognostic indicator in critical patients with chronic heart failure. ESC Heart Fail. 2022;9:1360–1369. doi: 10.1002/ehf2.13825

46. Wang Y. Stage 1 hypertension and risk of cardiovascular disease mortality in United States adults with or without diabetes. J Hypertens. 2022;40:794–803. doi: 10.1097/hjh.0000000000003080

47. Wang Y, Fang Y, Magliano DJ, Charchar FJ, Sobey CG, Drummond GR, Golledge J. Fasting triglycerides are positively associated with cardiovascular mortality risk in people with diabetes. Cardiovasc Res. 2023;119:826–834. doi: 10.1093/cvr/cvac124

